# Use and Perceptions of Artificial Intelligence Among Respondents to a Self-Selected Survey of Obstetrician-Gynecologists in France

**DOI:** 10.64898/2026.09.23.26363805

**Authors:** Romain Martineau, Clémentine Combes, Kevin Yauy, Philippe Deruelle

## Abstract

**Background:** Artificial intelligence (AI) is increasingly used in healthcare, but data on its real-world use and perceptions among obstetricians and gynecologists remain limited.

**Objectives:** To assess familiarity, use, perceptions, and expectations regarding AI among physicians practicing obstetrics and gynecology in France.

**Study Design:** We conducted a cross-sectional survey between June 2 and July 30, 2025, mainly distributed through professional organizations or associations and social media. Participation was voluntary and self-selected. AI use was defined as any use of an AI tool in routine clinical practice, regardless of type or frequency. Comparisons were made between AI users and non-users and between junior and senior physicians.

**Results:** Among 469 respondents, representing approximately 5% of the target population, 199 of 466 (42.7%) reported AI use in practice; three respondents were excluded from this analysis because of discordant questionnaire responses. AI use was associated with male gender and professional status, with residents more represented among users. Junior physicians reported greater AI use than senior physicians and expressed greater concern regarding medical confidentiality. Current applications primarily involved administrative tasks, medical writing, literature retrieval, and translation; anticipated future uses centered on clinical decision support and medical data interpretation. Perceptions of several AI applications differed between groups.

**Conclusions:** AI was reported to be used by a substantial proportion of respondents, primarily for supportive and informational tasks. These findings support further evaluation of training, validation, governance, and safe implementation of AI in obstetrics and gynecology.

**Highlights:**

- AI was used by 42.7% of respondents in routine clinical practice.
- AI use was more frequent among male and junior physicians.
- Current AI use mainly involved administrative and informational tasks.
- Seniority influenced acceptance of advanced clinical AI applications.
- Training, validation, and governance are key for safe AI implementation.

## Introduction

Artificial Intelligence (AI), encompassing machine learning and deep learning, enables complex data analysis and pattern recognition essential for modern healthcare [1–3]. In obstetrics and gynecology, AI has emerged as a promising tool for diagnostic imaging, surgical planning, and clinical decision support [4]. For obstetricians and gynecologists (Ob-Gyn), AI supports care from prenatal diagnosis to oncology and reproductive medicine. Its application across the care pathway has been accompanied by a substantial increase in research activity, with recent bibliometric analyses documenting a marked expansion of publications in obstetrics and gynecology [5].

Prenatal and intrapartum care represent major fields of AI application. AI enhances fetal imaging accuracy and aids prenatal diagnosis of congenital malformations, for example for facial profiles or cardiac assessments [6–8], and utilizes predictive models to stratify risks for major pregnancy complications, including preeclampsia, fetal growth restriction or preterm birth [9, 10]. During labor, AI-assisted electronic monitoring has been investigated for the prediction of neonatal acidemia and fetal asphyxia, although comparative performance relative to human judgment remains variable [11–14]. However, widespread adoption of these tools still requires rigorous clinical validation [15, 16].

Beyond obstetrics, AI supports gynecologic surgery through augmented reality, real-time navigation, and robotic integration, thereby improving operative outcomes (reduced complications and operative time) [17–19]. It also plays an emerging diagnostic and therapeutic role in urogynecology, benign disorders, and gynecologic oncology [20–23]. Furthermore, AI has been investigated for applications in assisted reproductive technologies [24] and medical education in obstetrics and gynecology [25].

Despite these promising advances, AI integration faces significant ethical and practical challenges, including algorithmic transparency, data confidentiality, and the crucial need to maintain human oversight [26–31]. The implementation of AI in clinical practice also raises questions regarding clinicians’ training and preparedness [32]. While many studies examine AI development and performance, data regarding real-world use and clinicians’ perceptions remain scarce [33–36].

To address this issue, this study aimed to describe familiarity, current practices, perceptions, and expectations regarding AI among respondents practicing obstetrics and gynecology in France.

## Materials and Methods

### Design and Setting

We conducted a descriptive cross-sectional survey using an electronic questionnaire to assess gynecologists’ and obstetricians’ perceptions, experiences, and expectations regarding AI. The survey was distributed between June 2 and July 30, 2025. The survey was distributed through multiple channels, including university and departmental hospitals, professional organizations, departmental councils, regional unions of health professionals, and social media platforms (LinkedIn, Facebook).

### Population and Criteria

Physicians were eligible if they were actively practicing gynecology and obstetrics in France during the study period. Those not practicing in the specialty or located outside France were excluded. Participants included residents, fellows, junior doctors, and senior specialists. Recruitment relied on multiple sources including professional associations, hospital departments, departmental boards, and social media. Because participation was voluntary and self-selected, representativeness of the target population could not be established.

The source population comprised physicians practicing obstetrics and gynecology in France. Because the survey was distributed through multiple professional and institutional channels and social media, no comprehensive sampling frame or list of invited physicians was available. The number of physicians who received or viewed the invitation was therefore unknown, and a response rate could not be calculated.

### Questionnaire

The questionnaire was designed to capture demographic information (gender, level of practice, type of institution, etc.), familiarity with AI, current applications of AI in clinical practice, perceived benefits and risks, and expectations for future AI integration and training. For the purposes of this survey, AI use was defined as any use of an artificial intelligence tool in routine clinical practice, regardless of the type of use or frequency of use. It included both open and closed-ended questions, but open-ended responses were not included in the present analysis. The questionnaire contained 40 questions. No pretest or pilot study was conducted. The full questionnaire is provided in S1.

### Data Collection Procedure

The questionnaire was administered electronically through email invitations, social media posts, and hospital departments. Participants provided electronic informed consent by checking a consent box before completing the questionnaire. Responses were collected without incentives. Email addresses were collected solely to prevent duplicate responses and were separated from survey responses before the research dataset was accessed. The analytical dataset therefore did not contain directly identifying information. However, no completely identical questionnaires were detected. Efforts were made to reach a broad sample of French gynecologists and obstetricians, with two follow-up emails from organizations that did not respond.

### Statistical Analysis

Categorical variables were summarized as frequencies and percentages. Likert-scale responses were initially coded into five categories and subsequently collapsed into two or three categories, depending on the item, by combining adjacent response options (e.g., very versus relatively negative/unfavorable responses, and very versus relatively positive/favorable responses). These recoding decisions were made after data collection and should therefore be considered post hoc. Although all survey questions were mandatory, responses considered inconsistent or discordant according to predefined criteria were excluded from the corresponding analyses, resulting in slight variations in the number of respondents across analyses.

Group comparisons were performed using the Chi-square test when its assumptions were met and Fisher’s exact test otherwise. Two prespecified comparisons were conducted: junior physicians (residents and junior attending physicians) versus senior physicians, and AI users versus non-users.

Variables associated with AI use in univariate analyses (p < 0.05) were entered into a multivariable logistic regression model to identify factors independently correlated with AI use and to characterize the profile of AI users.

Adjusted odds ratios (aORs) with 95% confidence intervals (95% CIs) were reported. Model calibration and goodness-of-fit were assessed using the Hosmer-Lemeshow test.

Two-sided p-values were reported for exploratory group comparisons, with p < 0.05 used as a conventional descriptive threshold. No adjustment for multiple comparisons was applied. Analyses were performed using SAS software. A study flow diagram is presented in Fig 1.

**Figure 1.**
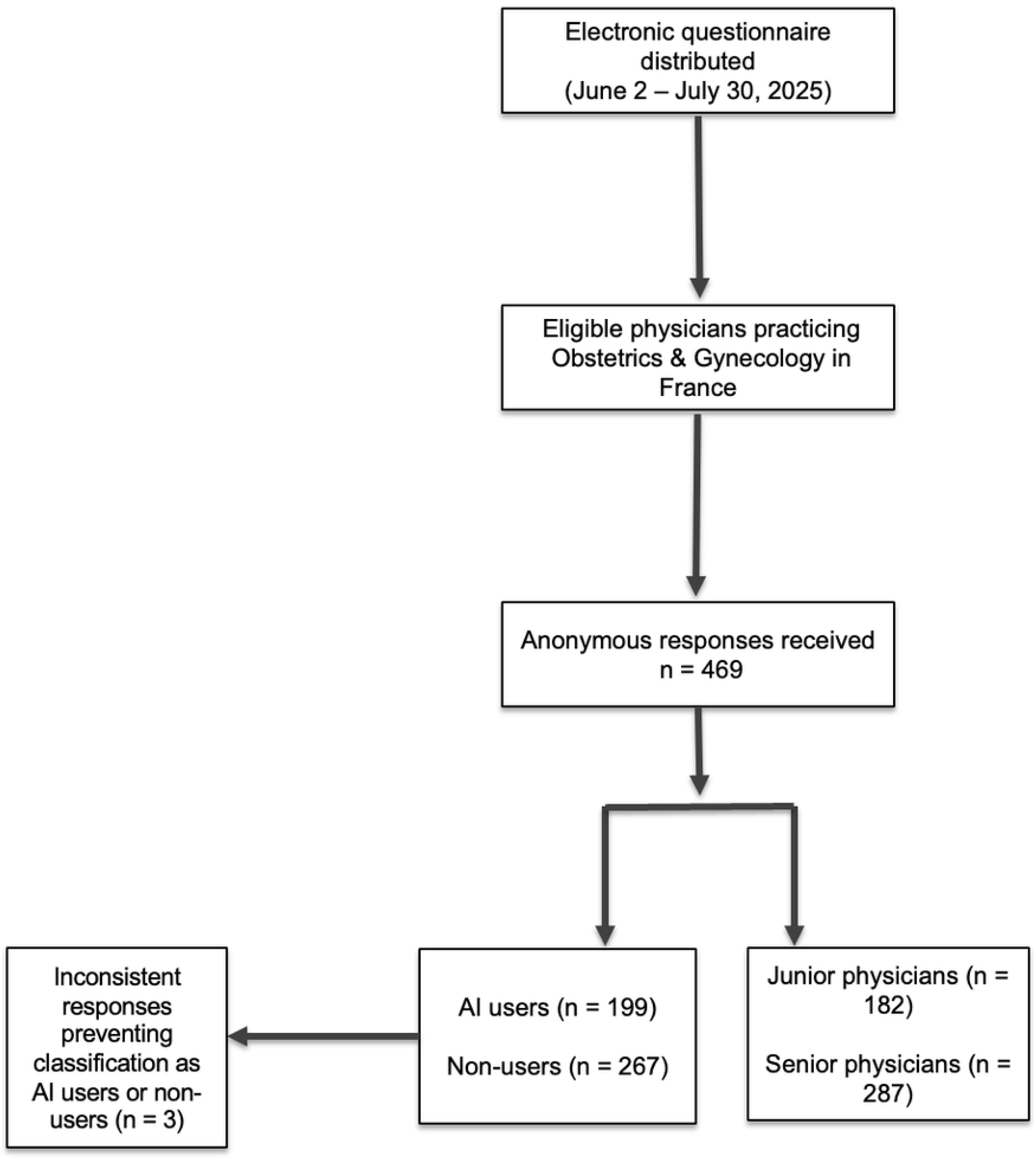
Study Flow Chart

### Ethical Considerations and Data Protection

Participation was voluntary and anonymous. Electronic informed consent to participate in the survey was obtained before questionnaire completion. Data were collected and stored in compliance with the General Data Protection Regulation (in French “RGPD”).

## Results

### Study Population (Table 1)

The 469 respondents practicing in France were predominantly female (77.2%, n = 362) and largely aged 25-34 years (49.5%, n = 232). Most were hospital or private practitioners (40.9%, n = 192) or residents (38.8%, n = 182), with the majority working in hospital settings (72%, n = 338) followed by private practice (17.1%, n = 80).

**Table 1.** Baseline Characteristics of the Study Population. Seperate file.

| Characteristics | Effective (n) | Rate (%) |
| --- | --- | --- |
| **Gender** |  |  |
| Female | 362 | 77.2 |
| Male | 106 | 22.6 |
| Non-binary | 1 | 0.2 |
| **Age** |  |  |
| < 25 years old | 11 | 2.35 |
| 25-34 years old | 232 | 49.5 |
| 35-44 years old | 116 | 24.7 |
| 45-54 years old | 53 | 11.3 |
| 55-65 years old | 38 | 8.1 |
| > 65 years old | 19 | 4.05 |
| **Qualification** |  |  |
| Residents | 182 | 38.8 |
| Clinical Fellows & Hospital Assistants | 42 | 9 |
| Hospital Consultants & Private Practitioners | 192 | 40.9 |
| Academic Physicians | 41 | 8.7 |
| Others | 12 | 2.6 |
| **Practice Setting** |  |  |
| Hospital-Based | 338 | 72 |
| Private Practice | 80 | 17.1 |
| Mixed | 39 | 8.3 |
| Private Clinic Employee | 8 | 1.7 |
| Other | 4 | 0.9 |
| **Additional Diploma** |  |  |
| First-Year Master's Degree | 132 | 28.1 |
| Second-Year Master's Degree | 78 | 16.6 |
| PhD Thesis | 52 | 11.1 |
| **Main Clinical Practice** |  |  |
| Medical Gynecology | 169 | 36 |
| Obstetrics | 234 | 49.9 |
| Benign Gynecological Surgery | 117 | 24.9 |
| Gynecological Oncological Surgery | 91 | 19.4 |
| Medical Assistance for Reproduction | 74 | 15.8 |
| Fetal Medicine | 55 | 11.7 |
| Abortion Care | 47 | 10 |
| Other | 24 | 5.1 |
| **Use of AI (n = 466)** |  |  |
| Yes | 199 | 42.7 |
| No | 267 | 57.3 |
| **Tools Used** |  |  |
| ChatGPT (Open AI) | 184 | 39.2 |
| Gemini (Google) | 17 | 3.6 |
| Copilot (Microsoft) | 20 | 4.3 |
| Le Chat Mistral AI | 21 | 4.5 |
| Apple Intelligence | 4 | 0.9 |
| DeepL AI | 25 | 5.3 |
| Other | 48 | 10.2 |
| **Mode of AI use in clinical practice**<br>**(among AI users, n = 199)** |  |  |
| Assistance with drafting letters/emails/summaries | 134 | 67.3 |
| Bibliographic research | 120 | 60.3 |
| Document / data summary (example: article summary) | 109 | 54.8 |
| Translation | 66 | 33.2 |
| Training / self-study | 41 | 20.6 |
| Therapeutic aid | 33 | 16.6 |
| Diagnostic aid | 31 | 15.6 |
| Informing a patient about their condition | 13 | 6.5 |

### Use of Artificial Intelligence (Table 2)

Among the 466 respondents with valid responses, 199 (42.7%) reported routine AI use; three respondents were excluded from this analysis because of discordant questionnaire responses. AI use differed according to several demographic and professional characteristics (Table 2, S2 Fig) : AI users were more frequently men (29.3% vs. 18%), aged 25–34 years (p = 0.02), and residents (p < 0.05). Furthermore, major differences emerged regarding perceived competence, training needs, and concerns over medical confidentiality or excessive AI reliance. Conversely, expectations regarding patient reactions did not differ between groups. AI use was associated with several demographic and professional characteristics (Table 2). The profile of AI users and non-users was illustrated using a radar chart (S2 Fig).

**Table 2.**
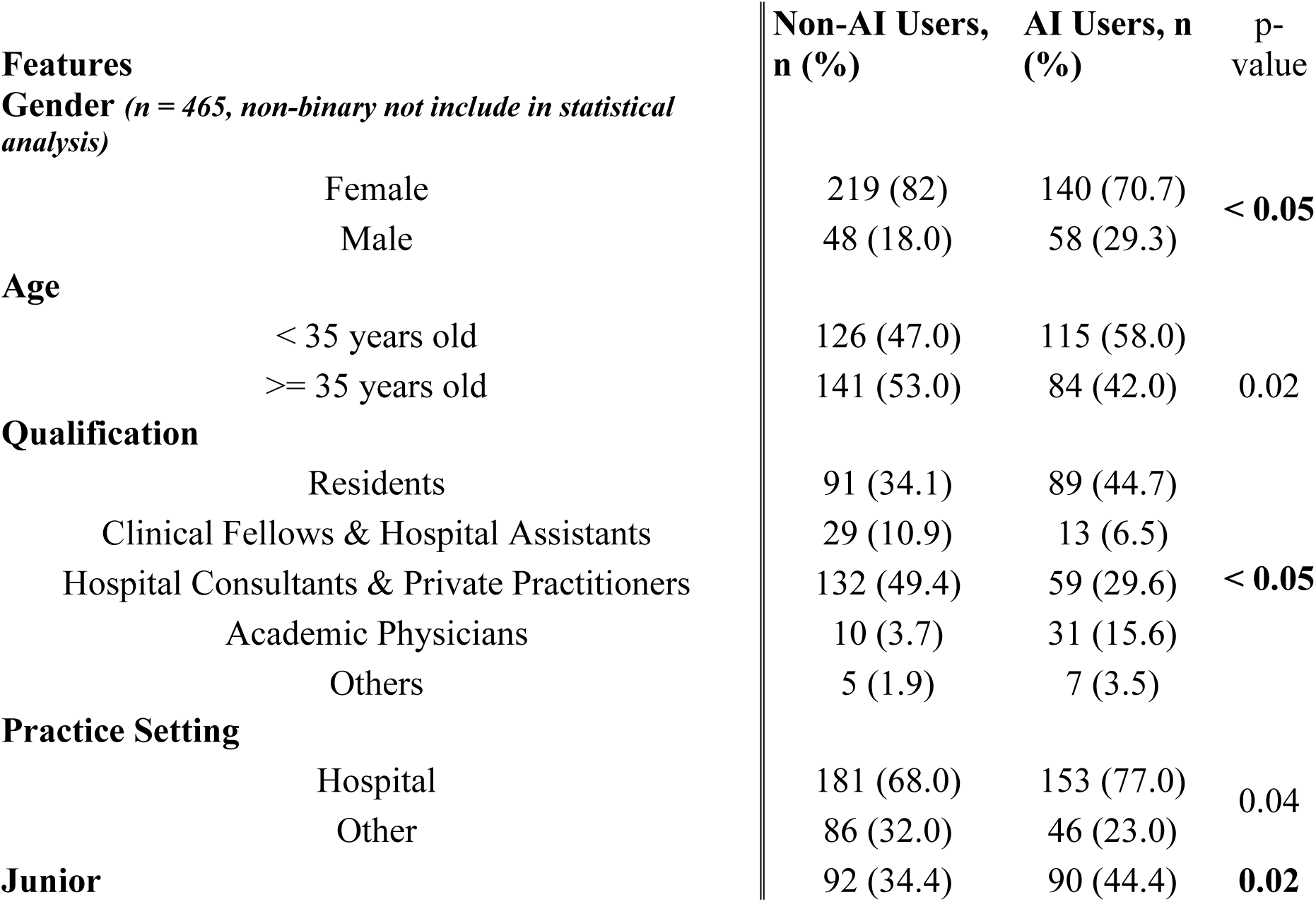
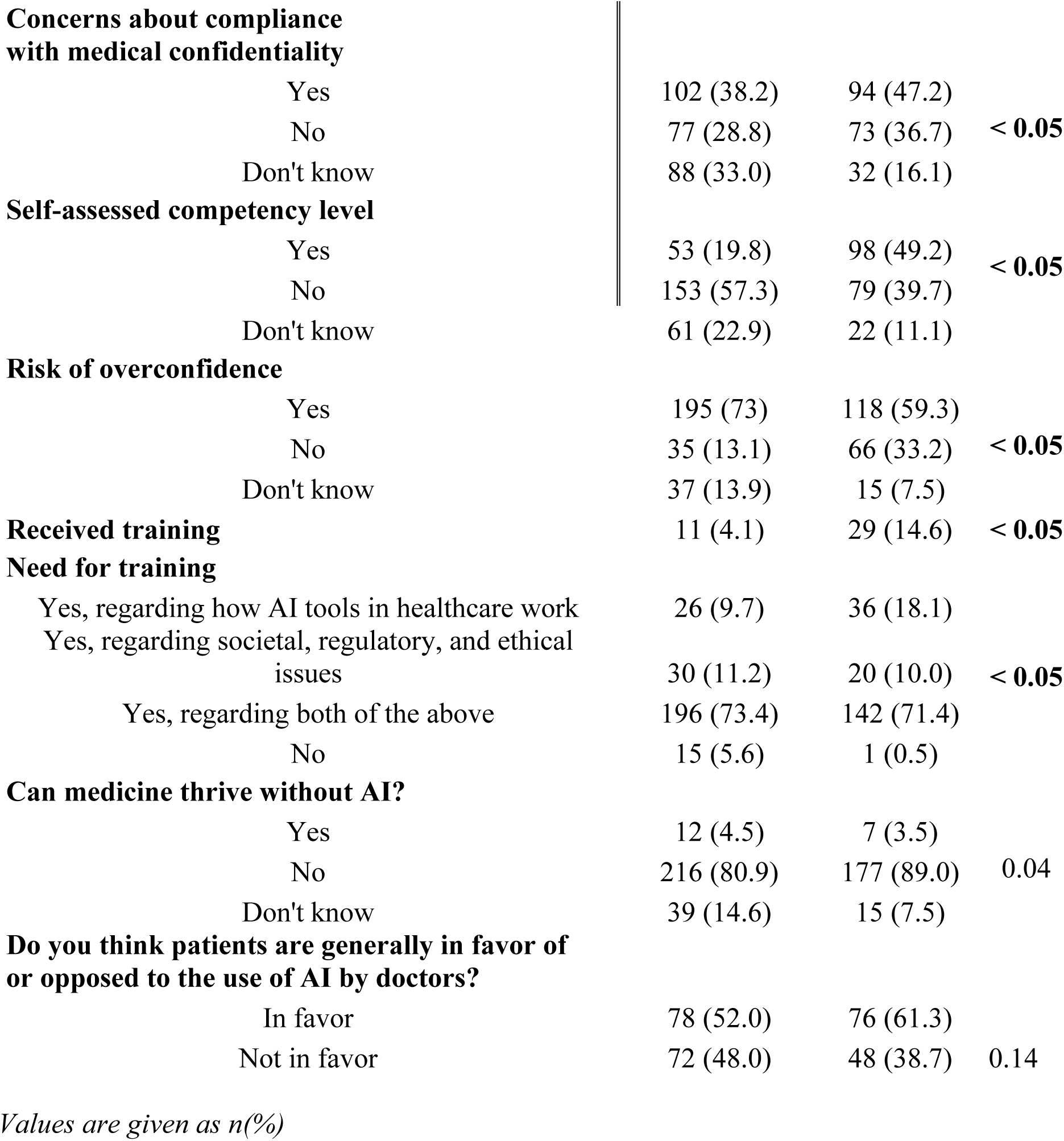
Comparison of Characteristics Between AI Users and Non-Users. Seperate file.

| Features |  | Non-AI Users, n (%) | AI Users, n (%) | p-value |
| --- | --- | --- | --- | --- |
| **Gender** ($n = 465$, non-binary not include in statistical analysis) | | | | |
|  | Female | 219 (82) | 140 (70.7) | < 0.05 |
|  | Male | 48 (18.0) | 58 (29.3) |  |
| **Age** |  |  |  |  |
|  | < 35 years old | 126 (47.0) | 115 (58.0) | 0.02 |
| | $\geq 35$ years old | 141 (53.0) | 84 (42.0) | |
| **Qualification** |  |  |  |  |
|  | Residents | 91 (34.1) | 89 (44.7) | < 0.05 |
|  | Clinical Fellows & Hospital Assistants | 29 (10.9) | 13 (6.5) |  |
|  | Hospital Consultants & Private Practitioners | 132 (49.4) | 59 (29.6) |  |
|  | Academic Physicians | 10 (3.7) | 31 (15.6) |  |
|  | Others | 5 (1.9) | 7 (3.5) |  |
| **Practice Setting** |  |  |  |  |
|  | Hospital | 181 (68.0) | 153 (77.0) | 0.04 |
|  | Other | 86 (32.0) | 46 (23.0) |  |
| **Junior** |  | 92 (34.4) | 90 (44.4) | 0.02 |

|  |  |  |  |
| --- | --- | --- | --- |
| **Concerns about compliance with medical confidentiality** |  |  |  |
| Yes | 102 (38.2) | 94 (47.2) |  |
| No | 77 (28.8) | 73 (36.7) | **< 0.05** |
| Don't know | 88 (33.0) | 32 (16.1) |  |
| **Self-assessed competency level** |  |  |  |
| Yes | 53 (19.8) | 98 (49.2) |  |
| No | 153 (57.3) | 79 (39.7) | **< 0.05** |
| Don't know | 61 (22.9) | 22 (11.1) |  |
| **Risk of overconfidence** |  |  |  |
| Yes | 195 (73) | 118 (59.3) |  |
| No | 35 (13.1) | 66 (33.2) | **< 0.05** |
| Don't know | 37 (13.9) | 15 (7.5) |  |
| **Received training** | 11 (4.1) | 29 (14.6) | **< 0.05** |
| **Need for training** |  |  |  |
| Yes, regarding how AI tools in healthcare work | 26 (9.7) | 36 (18.1) |  |
| Yes, regarding societal, regulatory, and ethical issues | 30 (11.2) | 20 (10.0) | **< 0.05** |
| Yes, regarding both of the above | 196 (73.4) | 142 (71.4) |  |
| No | 15 (5.6) | 1 (0.5) |  |
| **Can medicine thrive without AI?** |  |  |  |
| Yes | 12 (4.5) | 7 (3.5) |  |
| No | 216 (80.9) | 177 (89.0) | 0.04 |
| Don't know | 39 (14.6) | 15 (7.5) |  |
| **Do you think patients are generally in favor of or opposed to the use of AI by doctors?** |  |  |  |
| In favor | 78 (52.0) | 76 (61.3) |  |
| Not in favor | 72 (48.0) | 48 (38.7) | 0.14 |

*Values are given as n(%)*

Proportionally, men were more represented among AI users (29.3%) than among non-users (18%), in contrast to women. Age was also associated with AI use (p = 0.02), with the 25–34-year age group being the most prominent among users. Professional status also differed significantly between groups (p < 0.05), with residents being proportionally more prevalent among AI users. Differences were observed between groups in reported concerns regarding medical confidentiality, perceived competence, risk of excessive reliance on AI, and training. However, no difference was found regarding the perceived potential reaction of patients to the use of AI by healthcare professionals.

### Comparison Between Junior and Senior Practitioners (Table 3)

Junior practitioners were more frequently female than senior practitioners (90.1% vs. 69.2%, p < 0.05) and reported significantly greater current and potential use of AI. While both groups shared similar levels of perceived competence and training needs, juniors expressed greater concern regarding medical confidentiality breaches and were more likely to anticipate negative patient reactions to AI. The comparative analysis between junior and senior practitioners also revealed marked differences in demographic characteristics. Women were more represented in the junior group than in the senior group (90.1% vs 69.2%, p < 0.05). Junior practitioners reported significantly greater use of AI than senior practitioners. Practices and perceptions regarding artificial intelligence differed between the two groups, with junior practitioners more often reporting current or potential use of AI tools in their professional practice. Junior practitioners also appeared to express notably greater concern about potential breaches of medical confidentiality compared with senior practitioners. However, perceived competence in using AI, the risk of excessive reliance, the training received, and the perceived need for training did not differ significantly between groups. Furthermore, junior practitioners tended to believe that patients might have a more negative perception of AI use, whereas senior practitioners were more likely to consider that patients would view it positively.

**Table 3.**
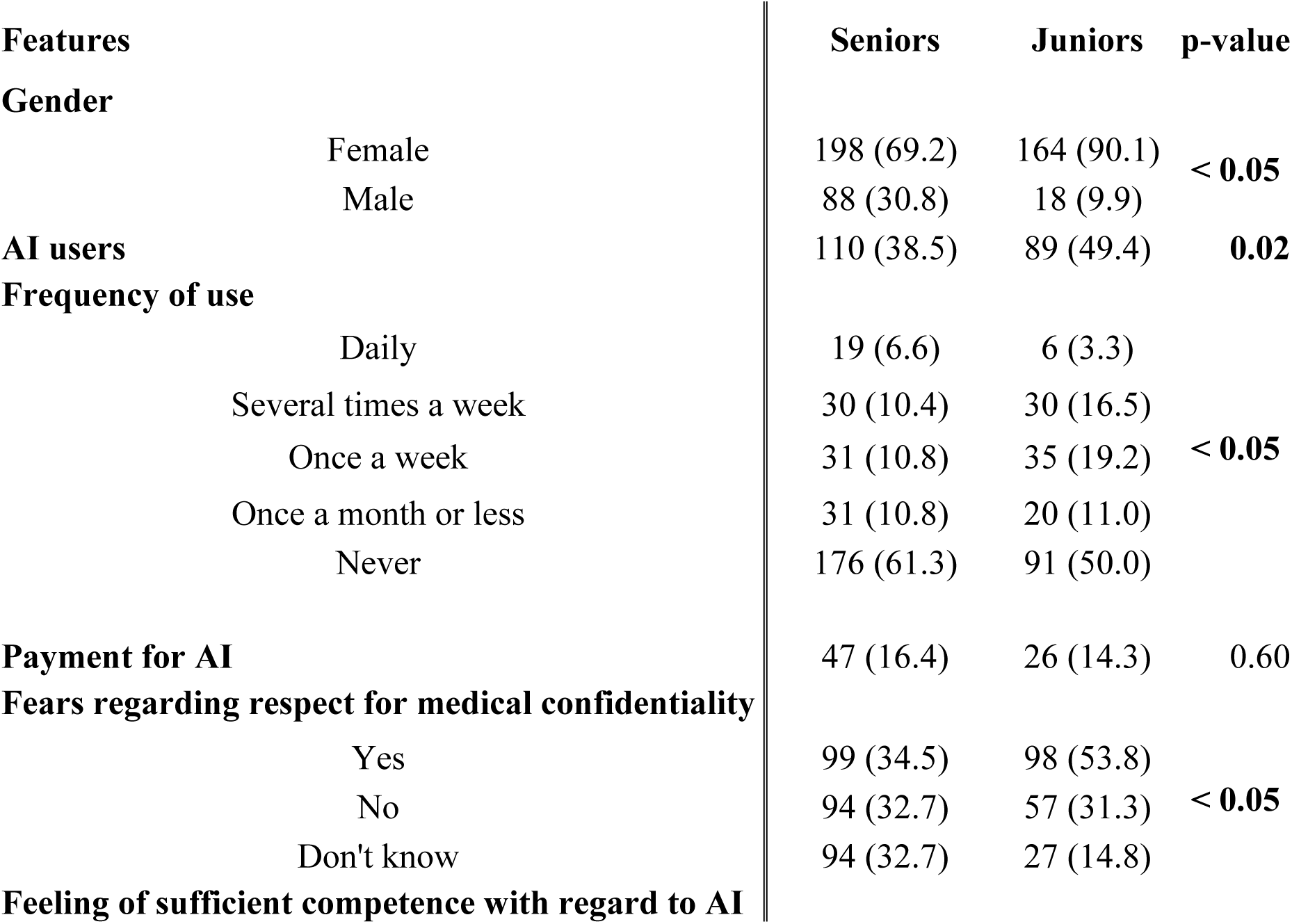
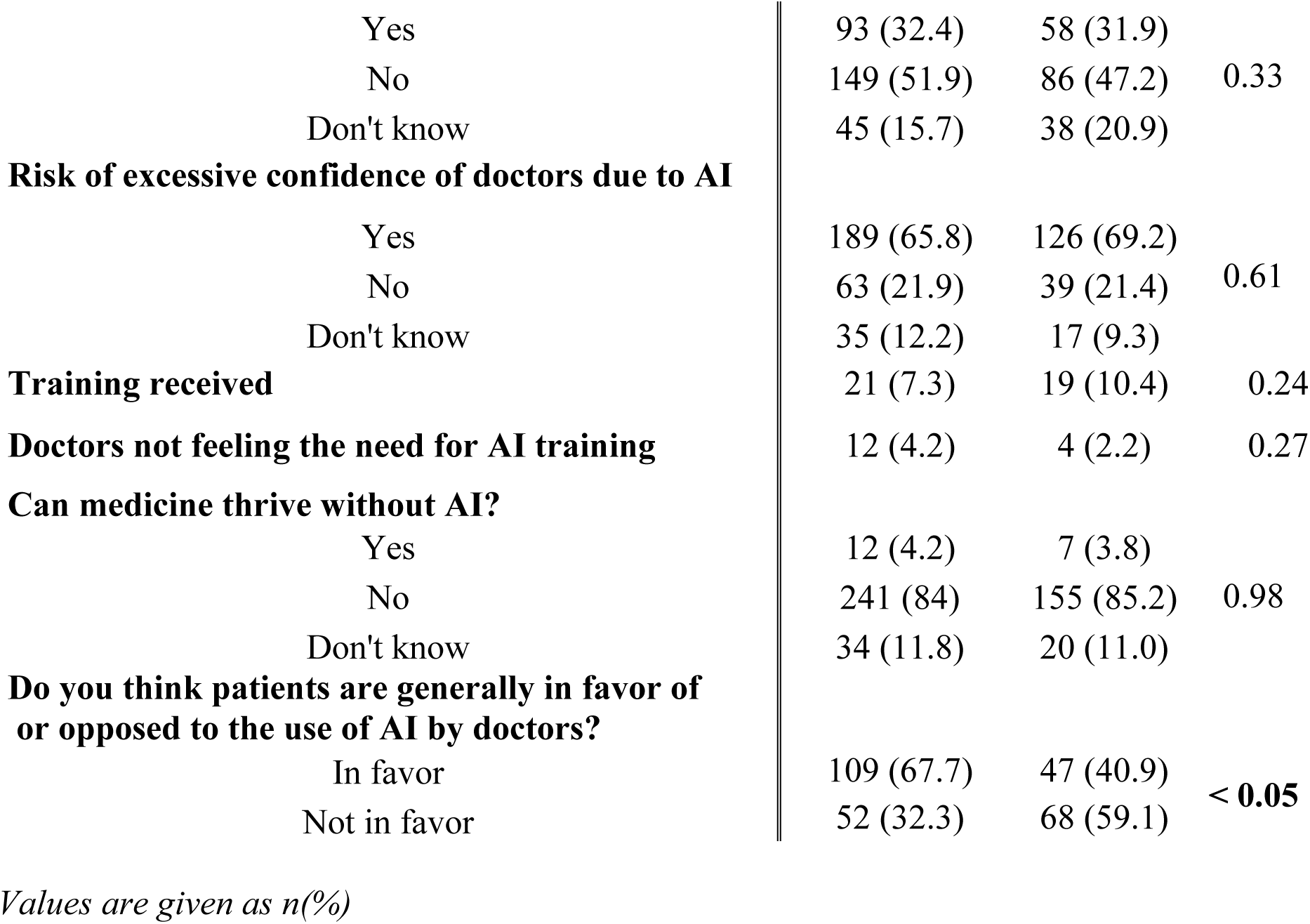
Comparison of Characteristics Between Junior and Senior Practitioners. Seperate file.

### Multivariable Analysis of Factors Associated with AI Use (Table 4)

In a multivariable logistic regression model (Hosmer-Lemeshow p = 0.973), only junior status (aOR 1.91, 95% CI 1.28–2.85, p = 0.001) and male gender (aOR 2.31, 95% CI 1.45–3.65, p < 0.001) were associated with AI use. Age and practice setting lost statistical significance after adjustment.

**Table 4.** Multivariable Analysis of Factors Associated With Artificial Intelligence Use. Seperate file.

Odds Ratio Estimates and Wald 95% Confidence Intervals
| Effect | Unit | Estimate | 95% Confidence Interval |  | p-value |
| --- | --- | --- | --- | --- | --- |
|  |  |  | Lower | Upper |  |
| Junior<br>(Yes vs.<br>No) | 1.0000 | 1.912 | 1.283 | 2.849 | 0.0014 |
| Gender<br>(Male vs.<br>Female) | 1.0000 | 2.305 | 1.454 | 3.652 | 0.0004 |

After adjustment, only professional seniority and gender remained independently correlated with AI use. Junior practitioners had higher odds of reported AI use than senior practitioners (aOR 1.91, 95% CI 1.28-2.85, p = 0.001). Male practitioners had higher odds of reported AI use than female practitioners (aOR 2.31, 95% CI 1.45-3.65, p < 0.001).

The Hosmer-Lemeshow goodness-of-fit test was not statistically significant (Chi2 = 0.001, degrees of freedom = 1, p = 0.973).

### Current and Potential Uses of Artificial Intelligence (Table 1 and 5)

Current AI applications primarily involve administrative tasks and medical writing, literature retrieval, and translation while anticipated future uses center on clinical decision support and medical data interpretation (**Table 1**).

Acceptability of specific AI applications varied significantly by usage status and professional seniority (**Table 5**). Compared to non-users, AI users held noticeably more favorable views regarding virtual assistants and AI’s potential to improve healthcare access in underserved areas (p < 0.05). They also perceived a more positive overall impact of AI on diagnostic performance, work quality, working time, and the physician-patient relationship (all p < 0.05). Although statistically significant, these differences were generally of moderate magnitude.

**Table 5.**
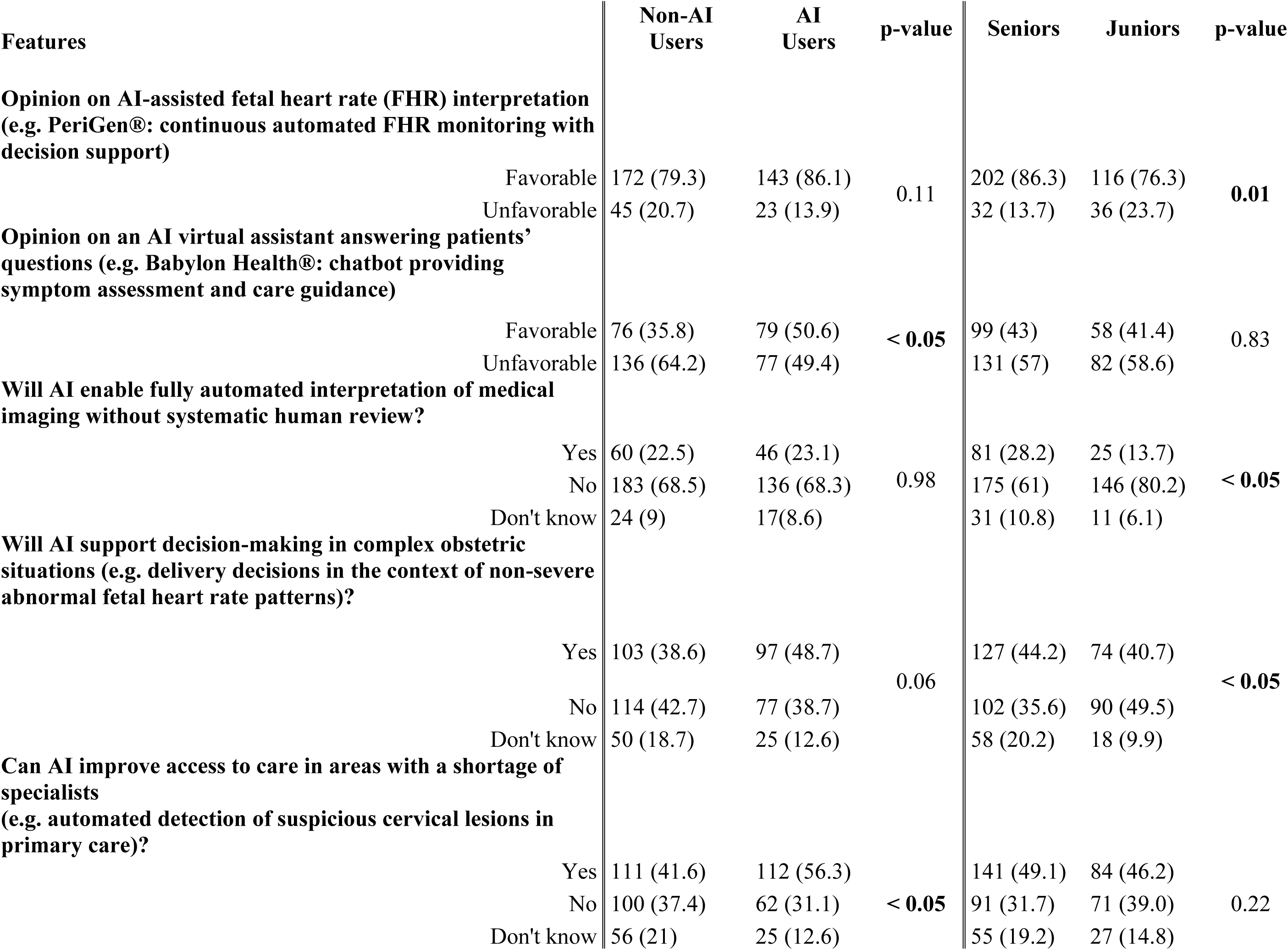
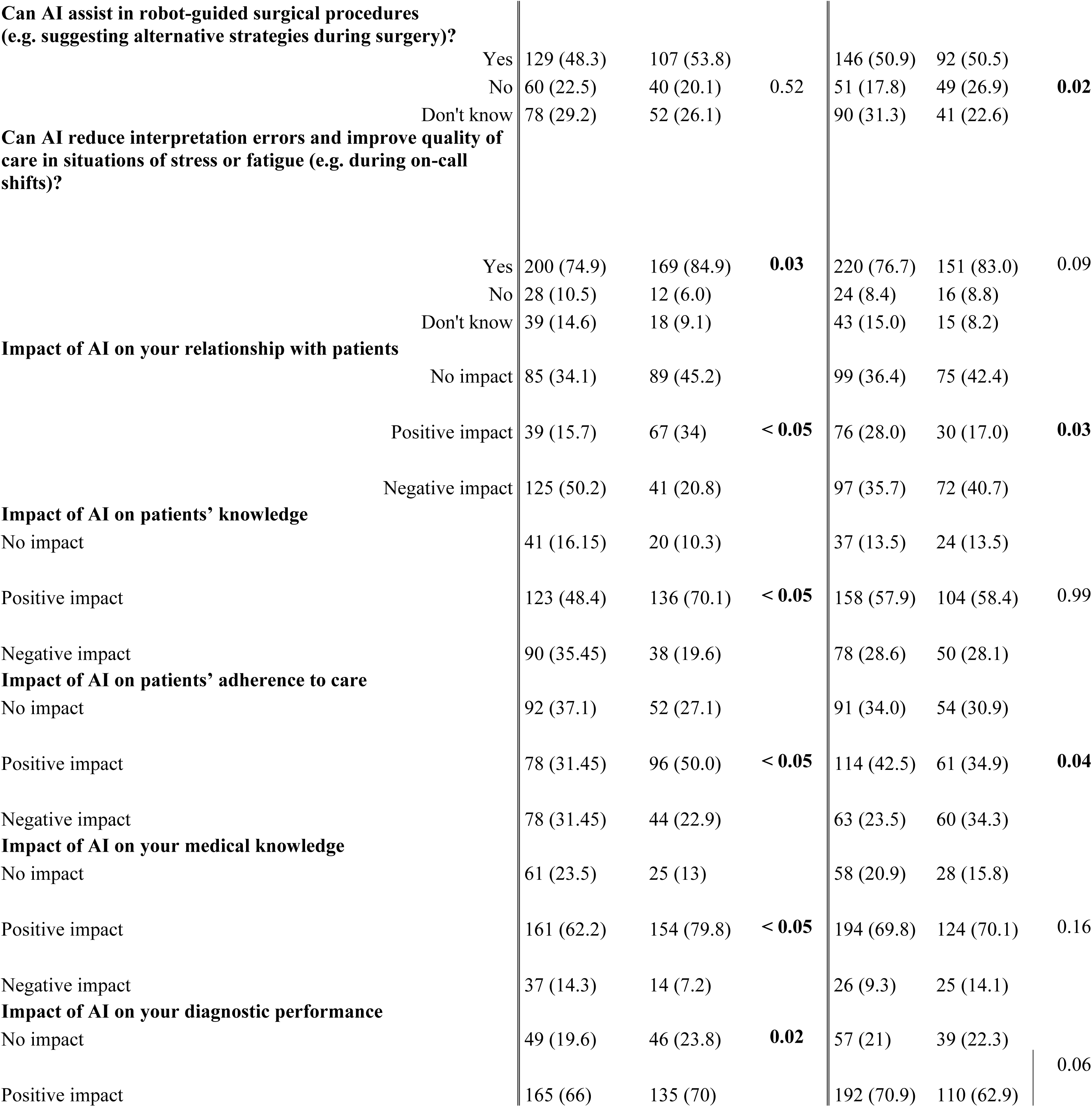
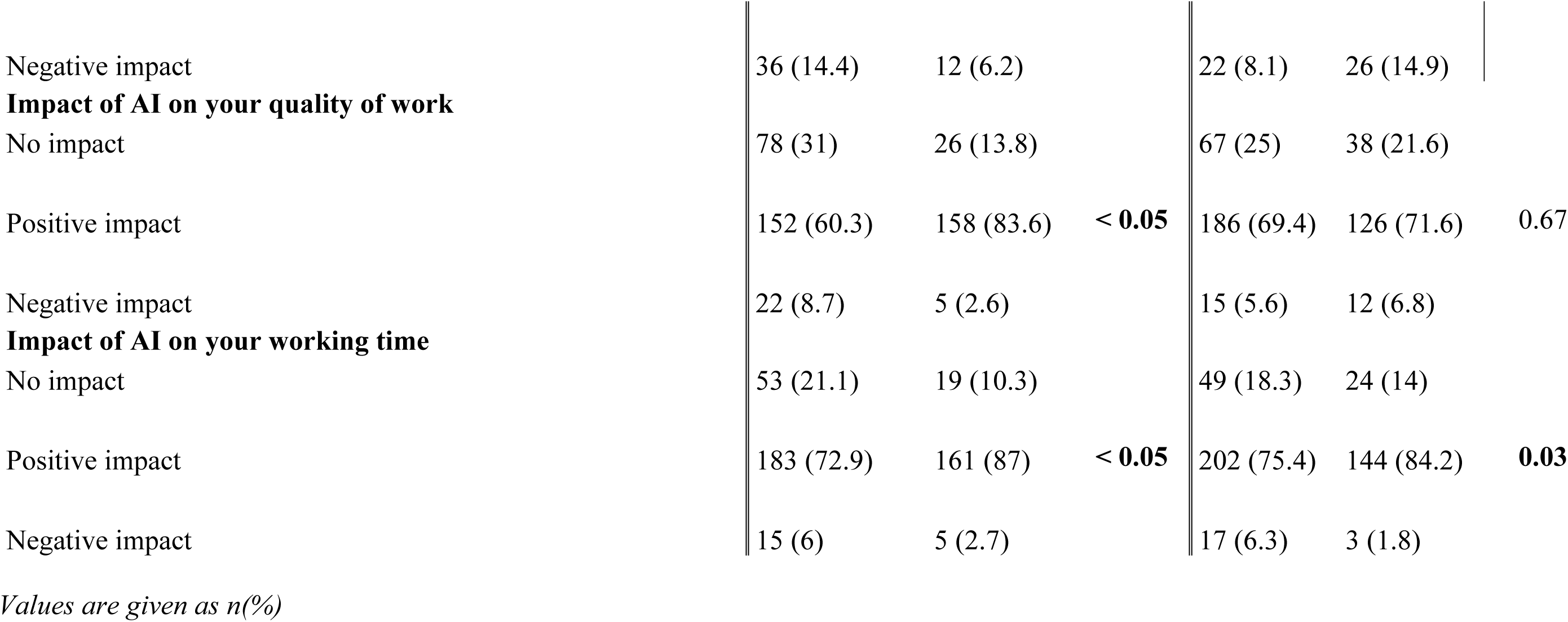
Current and Potential Applications of Artificial Intelligence According to AI Use Status and Practitioner Seniority. Seperate file.

In terms of seniority, senior physicians were significantly more receptive than juniors to advanced clinical applications, including AI-assisted fetal heart rate interpretation (p = 0.01), fully automated imaging evaluation, complex obstetric decision-making, and robotic surgery (all p < 0.05). Conversely, junior physicians were more likely to view AI primarily as a means to reduce working time (p = 0.03). Overall, prior AI use mainly influenced perceptions of AI’s organizational benefits, whereas professional seniority was more strongly associated with acceptance of advanced clinical applications (**Table 5**).

Detailed comparisons of non-significant parameters across groups are provided in Supplementary Material (S3 Table)

## Discussion

### Principal findings

This survey provides a contemporary exploratory description of AI use and perceptions among respondents practicing obstetrics and gynecology in France. Nearly half of respondents reported using AI tools in routine clinical practice, primarily for supportive tasks such as administrative and informational activities. AI use varied according to professional characteristics, suggesting heterogeneous adoption across practitioners. Although clinicians expressed interest in the potential benefits of AI, they also reported concerns regarding confidentiality, reliability, and professional responsibility, indicating a balanced perception of both the opportunities and the risks associated with AI implementation.

### Results in the context of what is known

The present findings are broadly consistent with previous studies conducted in the general medical population. The reported frequency of AI use in our sample falls within the range reported in previous international studies, although direct comparisons are limited by differences in populations, definitions of AI use, and study periods [37,38]. These differences likely reflect the rapid evolution of AI technologies, disparities in access to AI tools, and variations in professional environments.

Similarly, the predominance of administrative and informational applications observed in our study is in line with previous reports indicating that AI is currently used mainly to improve productivity, documentation, and information management rather than to support complex clinical decision-making [37,39]. The coexistence of enthusiasm and concern among respondents is also consistent with previous literature emphasizing that trust, transparency, and adequate validation remain key determinants of AI adoption in healthcare [37,38].

A major contribution of the present study relates to the specific context of obstetrics and gynecology. Although publications describing AI applications in this specialty have increased substantially in recent years, bibliometric analyses and systematic reviews have shown that the literature has primarily focused on algorithm development and technical performance, with limited evidence regarding clinical implementation and clinicians’ real-world practices [40–42]. Data specifically addressing obstetricians’ and gynecologists’ attitudes toward AI remain scarce. Existing studies are mainly qualitative or limited to specific clinical applications and consistently identify perceived usefulness, trust, validation, and explainability as major determinants of AI adoption [43]. Furthermore, recent evidence suggests that AI may support clinical decision-making in obstetrics and gynecology, including in time-sensitive clinical settings [44]. In this context, quantitative surveys focusing specifically on obstetricians and gynecologists remain limited. Our findings provide exploratory data describing reported AI use and perceptions among survey respondents within this specialty.

### Clinical implications

The heterogeneous patterns of reported AI use observed among respondents have several implications for clinical practice and healthcare systems. First, the variability in reported AI use suggests that further evaluation of clinicians’ training and educational needs may be warranted. Second, the coexistence of perceived benefits and concerns underscores the importance of establishing clear institutional and regulatory frameworks addressing data protection, clinical validation, and professional accountability before broader implementation. From an organizational perspective, the reported patterns of AI use suggest that respondents perceive potential effects on clinical workflow and physician workload. In particular, AI-assisted documentation and information retrieval may be perceived as potentially useful for reducing administrative burden and improving efficiency, although these effects were not directly evaluated in this survey [39]. In light of the reported patterns of use and perceptions, AI may currently be viewed as an assistive technology that complements rather than replaces clinicians’ expertise, particularly in a high-risk specialty such as obstetrics and gynecology. Additional prospective evaluation and robust regulatory oversight will be required before broader integration into routine clinical care.

### Research implications

Several questions remain unanswered. Future research should evaluate the clinical impact of AI tools in obstetrics and gynecology, particularly with respect to patient outcomes, safety, and quality of care. Additional studies are also needed to establish appropriate regulatory frameworks, better understand patients’ perspectives regarding AI use in women’s healthcare, and determine the most effective strategies for incorporating AI into both medical education and routine clinical practice. Longitudinal studies assessing changes in AI adoption over time and interventional studies evaluating the impact of education and governance strategies would further inform evidence-based implementation.

### Strengths and limitations

This study has several strengths. It provides exploratory data on AI use and perceptions among a relatively large sample of respondents practicing obstetrics and gynecology in France. Furthermore, the combined evaluation of both current practices and perceptions provides a comprehensive overview of clinicians’ engagement with AI tools in routine practice.

Several limitations should nevertheless be acknowledged. First, participation was voluntary and self-selected, and the survey was distributed through professional organizations, hospital departments, email, and social media. The number of physicians who received or viewed the invitation was unknown, and a response rate could therefore not be calculated. The respondents should consequently not be considered representative of all obstetricians and gynecologists practicing in France, and the observed proportions should not be interpreted as national prevalence estimates. Nevertheless, based on the approximate number of physicians currently in training and practicing in France (approximately 1,600 obstetrics and gynecology residents, 400 medical gynecology residents, 6,000 practicing obstetrician-gynecologists, and 1,400 practicing medical gynecologists [45–48]), our 469 respondents would correspond to an estimated response rate of approximately 5 %. Self-selection may have introduced selection bias, including overrepresentation of physicians with greater interest in artificial intelligence. Moreover, the relatively high proportion of younger physicians and residents among respondents may have influenced the observed estimates of AI use and perceptions. Second, the questionnaire was not pretested or piloted, and its measurement validity was not formally assessed. Third, some Likert-scale responses were collapsed and responses meeting predefined criteria for discordance were excluded from the corresponding analyses, which may have affected the observed group comparisons. Finally, because multiple comparisons were performed without adjustment for multiplicity, statistically significant findings should be interpreted as exploratory and may include false-positive findings.

## Conclusions

Among respondents to this self-selected survey, artificial intelligence was reported to be used by a substantial proportion of physicians, primarily for supportive and informational tasks. Although clinicians recognize its potential benefits, broader integration into clinical practice remains dependent on addressing key challenges related to trust, training, validation, confidentiality, and professional responsibility. These findings support further research into the training, governance, validation, and safe implementation of AI in obstetrics and gynecology, while providing a foundation for future research evaluating its clinical impact and optimal integration into routine care.

## Data Availability

De-identified data may be available from the corresponding author upon reasonable request, subject to applicable ethical and data-protection requirements.

## Acknowledgments

The Authors thank all the individuals who participated in the survey. The Authors also thank Elan Languages for providing English language editing.

## Presentation at a scientific meeting

This manuscript includes work that was previously presented as a poster at the “Paris Santé Femmes” Congress, Paris, France, December 2025.

## Author Contributions

All Authors: Conceptualization, data curation, formal analysis, methodology, validation; Writing – review and editing. R. Martineau and P. Deruelle: Supervision; Writing – original draft.

## Ethical Approval Statement

The authors attest that this is an original submission, it has not been previously published, nor submitted elsewhere, and that all authors have read and approved the submission.

## AI-use disclosure

Artificial intelligence was not used in the preparation of this manuscript.

## Supporting information

**S1 - Study Questionnaire**

Seperate file.

**S2 Fig - Radar Plot Comparing selected Characteristics of AI Users and Non-Users**

Characteristics of AI users are shown in blue and those of non-users in yellow.

Seperate file.

**S3 Table – Supplementary Table. Detailed results for variables with non-significant differences between AI users and non-users and between junior and senior practitioners.**

Seperate file.

